# Epidemiology of Cardiac Structure and Function Among African Caribbeans in Tobago: A Population-based Cohort Study

**DOI:** 10.64898/2026.11.18.26368418

**Authors:** Nirmalendu Deb Nath, Iva Miljkovic, Rain Katz, James Hosner, Ravinder Mallugari, Ryan Cvejkus, Lauren A Cruz, Victor Wheeler, Clifford N Thomas, Deepak K Gupta, Allison L Kuipers

## Abstract

**Background:** Individuals of African genetic ancestry are at high risk for heart failure (HF), which is often preceded by cardiac structural and functional abnormalities. Nevertheless, little is known about cardiac structure and function among Afro-Caribbeans residing in Tobago. This study aimed to investigate the epidemiology of cardiac structure and function among African Caribbeans in Tobago.

**Methods:** A total of 924 participants (41-83 years) underwent clinical evaluation and transthoracic echocardiography. Images were analyzed for left ventricular (LV) mass index (MI), left atrial volume index, diastolic dysfunction (DD), LV ejection fraction (LVEF), global longitudinal strain (GLS), E/e′ ratio, E/A ratio, and other echocardiographic measures. Cardiac structure and function were evaluated by age, sex, and HF stage.

**Results:** The mean age was 61.7 years, with 61.4% women, 47.8% obesity, and 20.2% diabetes. Average systolic blood pressure was approximately 140 mmHg, and 76.7% had hypertension. Abnormal LV geometry was present in 77.1% of participants, predominantly concentric remodeling (69.5%). The prevalence of DD was 13.4%, LVEF <50% was 1.5%, and abnormal GLS (≤-18%) was 5.2%. Women had significantly higher E/e′ ratios and a higher prevalence of Stage A and Stage B HF, whereas men had a higher prevalence of symptomatic (Stage C) HF. Participants aged 65-83 years had significantly higher E/e′ ratio, concentric remodeling, and LV hypertrophy than those aged 41-57 years.

**Conclusions:** Women and older adults have higher LV structural abnormalities and DD than men and younger adults. These results will be helpful to inform public health priorities related to cardiovascular health in Tobago.

## Introduction

Heart failure (HF) is a chronic condition and major public health problem affecting tens of millions of people worldwide. It is closely associated with other chronic diseases such as hypertension, atrial fibrillation, diabetes mellitus, obesity, and chronic kidney disease and carries an increased risk of death ^1^. In 2023, the prevalence of HF among African Americans was 3.6%, and age-adjusted mortality was 117.5 per 100,000 population ^2^. In addition, approximately 1.5 million African American adults have HF, and it is expected to increase by 29% by the year of 2030 ^3^. Similarly, cardiovascular disease (CVD), including HF, is also increasing in Caribbean Islands and it is one of the leading causes of morbidity and mortality ^4,5^. In 2021, the age-adjusted incidence and mortality of CVD was highest in Haiti among Caribbean nations, with 840 and 466 per 100,000 population, respectively ^6^.

Self-identified African Americans and Hispanic patients are at high risk of developing HF compared to other racial groups. Results from echocardiographic studies in the United States (U.S.) showed a 3% higher prevalence of HF in Black compared to White populations with Black individuals more likely to develop abnormal systolic function and left ventricular hypertrophy (LVH) ^7^. Similarly, an echocardiographic study of HF in the United Kingdom (U.K), including 211 Afro-Caribbean patients, reported that left ventricular septal wall thickness was significantly higher among African Caribbean patients compared their White counterparts ^8^. In high-income regions (e.g., North America and Europe), ischemic heart disease is the common cause of HF, and the prevalence of HF with preserved ejection fraction is becoming prevalent ^9^. In contrast, data from Africa and South Asia indicate that hypertension, cardiomyopathy, and rheumatic valvular disease remain major concern for structural heart abnormalities and HF ^10,11^. Although information of HF in Caribbean islands is limited, a study of 100 consecutive patients with symptomatic HF in Jamaica reported that hypertension was the most common cause of HF ^12^. Similarly, a study conducted by Yan et al. in Haiti identified hypertension and other chronic conditions, including chronic kidney disease and obesity, as major contributing factors for HF ^5^.

Despite advances in cardiac management and treatment, the risk of HF and other CVDs remains high in Caribbean regions including Tobago. One of the major risk factors for HF, CVD, and cardiac abnormalities is hypertension and its prevalence is increasing at an alarming rate among the people from African genetic ancestry throughout the world ^4,13^. Our preliminary cohort studies in Tobago reported a high prevalence (62%) of stage 2 hypertension, with the majority (78%) of cases not adequately controlled ^14^. Consequently, high blood pressure puts the African genetic ancestry individual of Tobago at increased risk of hemodynamic complications, such as arterial stiffness, LVH, and HF. However, no prospective studies have been conducted to study cardiac structure and function in Tobago. Echocardiography is a well-validated test and used widely clinically in Tobago to assess the cardiac structure and function. Therefore, the aim of this study was to characterize age and sex specific echocardiographic assessments of cardiac structure and function and their distribution across HF stages in a population-based study of African Caribbeans in Tobago.

## Methods

### Study population

Tobago Health Study (THS) began as a population based prospective cohort study of African ancestry men aged ≥40 years old in the Caribbean Island of Tobago, Trinidad and Tobago ^15^. Briefly, men were initially recruited from 1997 to 2003 for prostate cancer screening, with subsequent follow-up assessments of body composition and associated risk factors. To be eligible to enroll in the study, men had to be ambulatory, noninstitutionalized, and free from terminal illness.

The present echocardiography ancillary study was conducted within the THS cohort and included both men and women. Starting in 2018, Tobagonian women were recruited into the THS using the same eligibility criteria as the original cohort (i.e., ambulatory, noninstitutionalized, and free of terminal illness). From 2021, transthoracic echocardiography assessment was performed for both men and women in the THS to study cardiac structure and function. Population-based recruitment concluded in November 2024, with echocardiography data available for a total of 924 participants (357 men, 567 women) aged 41-83 years. Approval for the study was granted by the Institutional Review Boards of both the University of Pittsburgh and the Tobago Ministry of Health and Social Services. All participants provided written informed consent prior to enrollment.

### Transthoracic echocardiography

Transthoracic echocardiography (TTE) was performed using a GE Vivid E9 ultrasound machine (GE Healthcare, Chicago, IL, USA) by trained sonographers in Tobago and under the supervision of board-certified cardiologists at Premier Heart Care Ltd from Port of Spain. Comprehensive 2-dimensionsal (2D), M-mode, and Doppler echocardiography were performed according to recommended American Society of Echocardiography (ASE) guidelines. Images were obtained using standard parasternal, apical, subcostal, and suprasternal views. Each 2D, M-mode, or Doppler image captured at least 3 full cardiac cycles in sinus rhythm and 5 in atrial fibrillation. Frame rates were maintained between 50-90 frames per second. Scales and baselines were adjusted as appropriate to capture peak inflow and/or regurgitant velocities. Attention was given to the electrocardiographic rhythm to avoid capturing images with ectopic beats. Echocardiography images were then processed through a central reading laboratory at Vanderbilt University Medical Center to abstract analytic variables using protocols similar to other prominent US population-based cohorts that include echocardiography ^16,17^. The core laboratory demonstrated high measurement reproducibility, with intra- and inter-reader intraclass correlation coefficients above 0.90 for left ventricular (LV) wall thickness, chamber dimensions, LV fraction (LVEF), and global longitudinal strain (GLS).

Analytical variables including LV Mass Index (LVMI), LV concentric remodeling, LVH, Left Atrium Volume Index (LAVI), E/A ratio, E/e’ ratio, diastolic dysfunction (DD), LV End Diastolic Volume (LVEDV), LV End Systolic Volume (LVESV), LVEF, GLS etc. were used in this study. Left ventricular mass and relative wall thickness (RWT) were used to classify subjects as normal, concentric remodeling, concentric hypertrophy and eccentric hypertrophy according to ASE 2015 chamber quantification guidelines ^18^. The LVRWT was calculated as twice the LV posterior wall thickness at end-diastole divided by the LV internal diameter at end-diastole with a value ≤0.42 considered normal. Patterns of LV geometric changes were classified as normal (normal LVMI and normal RWT), concentric remodeling (normal LVMI and increased RWT), concentric hypertrophy (increased LVMI and increased RWT) and eccentric hypertrophy (increased LVMI and normal RWT). In addition, LV diastolic dysfunction was classified as normal, Grade 1, grade 2, grade 3 ^18^.

### Covariate measurements

Blood pressure readings were taken three times while seated using an automated blood pressure cuff (Omron Healthcare, Inc, Vernon Hills, IL) with 5 minutes of rest in between measurements. The average of the 2nd and 3rd readings was used as the mean systolic and diastolic blood pressure (SBP and DBP), respectively. Hypertension was defined as SBP ≥ 130 mmHg, DBP ≥ 80 mmHg, or current use of antihypertension medication. Diabetes status was self-reported and/or based on medication inventory. Standing height was measured twice using a wall-mounted stadiometer and averaged to the nearest 0.1 cm. Weight was measured to the nearest 0.1 kg using a balance beam scale. Body mass index (BMI) was calculated as the ratio of weight to height squared (kg/m^2^). Standardized interviewer-administered questionnaires were used to collect demographic, health history, and lifestyle information, including smoking status, and sedentary behavior. Smoking was defined as current smokers, ex-smokers and never smokers. Sedentary behavior was characterized based on reported time spent watching television per week ^14,19^. All medications were recorded by staff through verification of pill bottles. Additionally, participants were categorized according to the American Heart Association and the American College of Cardiology stages of HF (A through C), with cardiac structure and function analyzed across these stages ^20^. Accordingly, participants with hypertension, CVDs, diabetes, obesity, and family history cardiomyopathy but normal cardiac structure and function were categorized as Stage A HF. The persons who had LVH (concentric or eccentric hypertrophy), valvular heart disease, reduced left ventricular or right ventricular systolic function, increased filling pressure, and abnormal DD were classified as Stage B HF. Finally, participants with current or prior symptoms and/or signs of HF with structural and functional cardiac abnormality were characterized as Stage C HF.

### Statistical analysis

All analyses were conducted in R statistical software version 4.2.2 ^21^. All continuous measures were assessed for normality. Normal distribution was evaluated using the Shapiro Wilk test, qq plot, and histograms. Mean, standard deviation, medians and interquartile ranges were used for summary statistics. In addition, box plots were also used to study the descriptive statistics of echocardiographic measures.

Sex stratified analysis was conducted to estimate the association of cardiac structure and function in men and women. In addition, data were stratified according to tertile of age, and the results were presented for 1^st^ tertile (age 41-57), 2^nd^ tertile (58-64), and 3^rd^ tertile (65-83) aged people to identify the association between age and echocardiographic outcomes. Sex and age differences were tested using t-tests, chi square test, and logistic regression as appropriate. For all models p < 0.05 were reported for significant associations.

## Results

A total of 924 African Caribbean adults were included in this Study (Table 1). Average age of this cohort was 60.7 years (range 41-83 years) with 61.4% female and 38.6% male. The study population had a mean BMI of 30.2 kg/m^2^, where female participants had a significantly higher BMI than male participants, with mean BMIs of 31.6 kg/m^2^ and 28.0 kg/m^2^ respectively. More than half of the participants were sedentary (50.9% watch >14 hours of TV/week). Although women were more sedentary than men, the percentage of current smokers was significantly higher in men compared to women. The mean SBP and DBP were approximately 140 mmHg and 79 mmHg, respectively, with a high prevalence (76.7%) of hypertension. The prevalence of hypertension was slightly higher in women (78.5%) than men (73.9%); but it was not statistically significant (p=0.119). Prevalence of obesity (47.8%) and self-reported diabetes mellitus (20.2%) was high, but history of pre-existing CVD was low (myocardial infarction 2.7%, stroke 1.7%, and atrial fibrillation 0.6%). Although no difference was observed between men and women for history of myocardial infarction and stroke, women had higher prevalence of a history of atrial fibrillation compared to men (0.7% versus 0.3%, respectively, p<0.001).

**Table 1:** Characteristics of men and women participants in the THS.

| **Characteristics [mean (SD) or %, as appropriate]** | **Overall<br>N=924** | **Men<br>N=357 (38.6%)** | **Women<br>N=567 (61.4%)** | **P-value<sup>1</sup>** |
|---|---|---|---|---|
| Age | 60.7 (6.8) | 61.7 (6.3) | 60.1 (7.0) | <0.001 |
| Body Mass Index (kg/m<sup>2</sup>) | 30.2 (5.9) | 28.0 (4.4) | 31.6 (6.2) | <0.001 |
| Obesity (%) | 47.8 | 30.8 | 58.6 | <0.001 |
| Systolic blood pressure (mmHg) | 139.6 (20.3) | 140.0 (20.2) | 139.0 (20.4) | 0.649 |
| Diastolic blood pressure (mmHg) | 79.2 (11.0) | 78.8 (11.7) | 79.5 (10.6) | 0.367 |
| Hypertension (%) | 76.7 | 73.9 | 78.5 | 0.119 |
| Diabetes (%) | 20.2 | 19.0 | 21.0 | <0.001 |
| Current smoking (%) | 3.7 | 7.6 | 1.2 | <0.001 |
| Watch TV>14hr/week (%) | 50.9 | 45.8 | 54.2 | 0.029 |
| Myocardial infarction (%) | 2.7 | 2.8 | 2.7 | 0.723 |
| Stroke (%) | 1.7 | 1.7 | 1.8 | 0.529 |
| Atrial fibrillation (%) | 0.6 | 0.3 | 0.7 | <0.001 |

SD=Standard Deviation.
<sup>1</sup>t-test for continuous outcomes and chi-square test for dichotomous outcomes were used to calculate the p-value.

Echocardiographic characteristics of the THS are shown in Table 2. Structural abnormalities were common among participants, with concentric remodeling 69.5%, concentric hypertrophy 6.1% and eccentric hypertrophy 1.5% (i.e., LVH in 7.6%). In addition, the mean aortic valve area was 2.5 cm^2^ and mean peak aortic valve velocity was 1.3 m/s. The prevalence of aortic stenosis of any severity was 2.4% in this THS. Average LVEF was 63.8% (range 25.3-74.4%), with 86.6% 11.9% 0.8%, and 0.8% of individuals displaying LVEF ≥60%, 50-59.9%, 40-49.9%, and <40% respectively. The markers of subclinical systolic dysfunction by GLS were present with 5.2% having GLS worse than -18% (4% having -16% to -17.7%, and 1.2% having >-16%), and mean tissue Doppler s wave velocity was 0.08 m/s. Average LVEDV and LVESV were 70.2 mL and 25.6 mL, respectively. The prevalence of any DD was 13.4%. Mean lateral and septal e’ velocities were 0.09 m/s and 0.07 m/s respectively. Mean LAVI was 24.0 ml/m^2^, while average Tricuspid Regurgitation velocity was 2.2 m/s. The mean value of E/e′ ratio was 9.2, with 37.9% of participants having a normal E/e′ ratio (<8.0), 57.5% intermediate E/e′ ratio (>8 to ≤14), and 4.6% had an elevated E/e′ ratio (>14).

**Table 2:** Cardiac structure and function assessed by echocardiography in the THS.

| Measures | Mean (SD),<br>or % | Median | IQR | Minimum | Maximum |
|---|---|---|---|---|---|
| ***LV structure*** |  |  |  |  |  |
| LV internal diameter diastole (mm) | 42.7 (4.9) | 42.4 | 6.6 | 27.8 | 69.0 |
| LV internal diameter systole (mm) | 27.4 (4.1) | 27.1 | 5.0 | 17.8 | 61.2 |
| LVEDV (ml) | 70.2 (16.3) | 67.9 | 19.1 | 24.8 | 182.9 |
| LVEDV Index to BSA (ml/m<sup>2</sup>) | 36.6 (7.9) | 35.7 | 9.6 | 13.8 | 94.3 |
| LVESV (ml) | 25.6 (8.9) | 24.1 | 8.2 | 7.3 | 136.6 |
| LVESV Index to BSA (ml/m<sup>2</sup>) | 13.3 (4.3) | 12.7 | 4.1 | 5.1 | 70.4 |
| LV septal wall thickness diastole (mm) | 10.7 (1.4) | 10.6 | 1.9 | 6.4 | 16.4 |
| LV posterior wall thickness diastole (mm) | 10.2 (1.4) | 10.1 | 1.8 | 6.6 | 16.1 |
| LV septal wall thickness systole | 14.2 (1.8) | 14.2 | 2.1 | 9.2 | 35.1 |
| LV posterior wall thickness systole | 14.0 (1.6) | 13.9 | 1.9 | 9.2 | 25.4 |
| LV relative wall thickness (%) | 48.4 (8.8) | 47.1 | 10.9 | 29.3 | 97.0 |
| LV mass (g) | 151.7 (39.5) | 146.3 | 49.6 | 63.1 | 333.9 |
| LVMI to BSA (g/m2) | 78.7 (18.1) | 76.3 | 22.1 | 40.1 | 167.5 |
| Normal geometry (%) | 22.8 | -- | -- | -- | -- |
| Concentric remodeling (%) | 69.5 | -- | -- | -- | -- |
| Concentric LVH (%) | 6.1 | -- | -- | -- | -- |
| Eccentric LVH (%) | 1.5 | -- | -- | -- | -- |
| ***LV function*** |  |  |  |  |  |
| LV outflow VTI (cm) | 22.6 (4.6) | 22.3 | 5.8 | 11.3 | 55.2 |
| Stroke volume (ml) | 67.7 (15.4) | 65.8 | 18.8 | 30.7 | 155.3 |
| Stroke volume index to BSA (ml/m<sup>2</sup>) | 35.3 (7.9) | 34.4 | 9.8 | 16.7 | 84.5 |
| Ejection fraction (%) | 63.8 (4.9) | 64.4 | 4.6 | 25.3 | 74.4 |
| Ejection fraction categories |  |  |  |  |  |
| ≥60% | 86.6 | -- | -- | -- | -- |
| 50%-59.9% | 11.9 | -- | -- | -- | -- |
| 40%-49.9% | 0.8 | -- | -- | -- | -- |
| <40% | 0.8 | -- | -- | -- | -- |
| LV TDI s’ lateral (m/s) | 0.08 (0.02) | 0.08 | 0.03 | 0.03 | 0.16 |
| GLS (%) | -22.1 (2.7) | -22.2 | 3.8 | -29.3 | -11.3 |
| GLS categories |  |  |  |  |  |
| Normal (≤-18%) | 94.8 | -- | -- | -- | -- |
| Mildly impaired (-16%-17.9%) | 4.0 | -- | -- | -- | -- |
| Severely impaired (>-16%) | 1.2 | -- | -- | -- | -- |
| LV TDI e’ lateral (m/s) | 0.09 (0.03) | 0.09 | 0.04 | 0.03 | 0.20 |
| LV TDI e’ septal (m/s) | 0.07 (0.02) | 0.07 | 0.03 | 0.02 | 0.14 |
| LV TDI A’ lateral (m/s) | 0.10 (0.02) | 0.09 | 0.03 | 0.04 | 0.19 |
| LV TDI A’ septal (m/s) | 0.10 (0.02) | 0.09 | 0.02 | 0.03 | 0.17 |
| Diastolic dysfunction (%) |  |  |  |  |  |
| None | 86.6 | -- | -- | -- | -- |
| Grade 1 | 11.5 | -- | -- | -- | -- |
| Grade 2 | 1.6 | -- | -- | -- | -- |
| Grade 3 | 0.3 | -- | -- | -- | -- |
| ***RV structure*** |  |  |  |  |  |
| RV diameter at base (mm) | 34.3 (4.3) | 34.0 | 5.8 | 23.8 | 58.7 |
| RV end diastolic area (cm<sup>2</sup>) | 13.9 (3.1) | 13.5 | 4.2 | 7.6 | 34.8 |
| RV end systolic area (cm<sup>2</sup>) | 7.6 (2.0) | 7.4 | 2.4 | 2.3 | 28.3 |
| ***RV function*** |  |  |  |  |  |
| RV fractional area change (%) | 45.0 (4.9) | 45.4 | 5.9 | 18.7 | 86.1 |
| TAPSE (mm) | 21.9 (2.9) | 21.6 | 4.0 | 14.5 | 31.3 |
| S’ (m/s) | 0.14 (0.02) | 0.13 | 0.03 | 0.06 | 0.22 |
| RVOT VTI | 16.4 (3.4) | 16.2 | 4.4 | 6.2 | 30.8 |
| ***LA structure and function*** |  |  |  |  |  |
| LA diameter (mm) | 32.3 (4.1) | 31.9 | 5.0 | 23.0 | 58.0 |
| LA volume (ml) | 46.2 (13.8) | 44.0 | 16.3 | 15.7 | 151.2 |
| LAVI to BSA (ml/m<sup>2</sup>) | 24.0 (7.0) | 23.2 | 8.2 | 8.4 | 77.9 |
| ***Mitral valve*** |  |  |  |  |  |
| Peak E velocity (m/s) | 1.3 (0.3) | 1.3 | 0.3 | 0.7 | 4.7 |
| Peak A velocity (m/s) | 0.8 (0.2) | 0.8 | 0.2 | 0.4 | 1.4 |
| E/A ratio | 0.9 (0.2) | 0.8 | 0.3 | 0.4 | 2.3 |
| ***Aortic valve*** |  |  |  |  |  |
| Peak velocity (m/s) | 1.3 (0.3) | 1.3 | 0.3 | 0.7 | 4.7 |
| Mean gradient (mm Hg) | 19.5 (5.4) | 19.3 | 6.6 | 4.0 | 46.5 |
| Aortic valve area (cm<sup>2</sup>) | 2.5 (0.6) | 2.5 | 0.7 | 0.6 | 5.0 |
| Aortic stenosis |  |  |  |  |  |
| No (%) | 97.6 | -- | -- | -- | -- |
| Yes (%) | 2.4 | -- | -- | -- | -- |
| ***Hemodynamics*** |  |  |  |  |  |
| Cardiac output (l/min) | 4.6 (1.2) | 4.5 | 1.4 | 1.8 | 11.8 |
| Cardiac index to BSA (l/min/m<sup>2</sup>) | 2.4 (0.6) | 2.3 | 0.7 | 0.8 | 5.8 |
| Average E/e’(ratio) | 9.2 (2.8) | 8.8 | 3.1 | 4.3 | 30.2 |
| TR velocity (m/s) | 2.2 (0.3) | 2.2 | 0.4 | 1.0 | 3.4 |
| PASP (mm Hg) | 24.5 (5.4) | 24.3 | 6.7 | 9.0 | 51.5 |
| Pulmonary vascular resistance (WU) | 1.5 (0.3) | 1.5 | 0.4 | 0.6 | 3.6 |

SD=Standard Deviation. IQR=Interquartile Range; LV=Left Ventricular; LVEDV=Left Ventricular End Diastolic Volume; BSA=Body Surface Area; LVMI= Left Ventricular Mass Index; LA=Left Atrium; LAVI= Left Atrium Volume Index; RV=Right Ventricular; TAPSE=Tricuspid Annular Plane Systolic Excursion; GLS=Global Longitudinal Strain; RVOT VTI=Right Ventricular Outflow Tract Velocity-Time Integral; TDI=Tissue Doppler Imaging; TR= Tricuspid Regurgitation; PASP= Pulmonary Artery Systolic Pressure. It was calculated using TR velocity and right atrial pressure of 5 mm Hg.

Sex and age specific cardiac structure and function metrics are shown in Table 3, Table 4, as well as Supplemental table 1. Women demonstrated a higher LVEF than men (64.1% vs. 63.4%, p = 0.021) (Figure 1A), whereas men had significantly higher LVEDV (76.0 vs 66.6 mL, p<0.001), LVESV (28.2 vs. 24.0 mL, p < 0.001), and stroke volume (70.6 vs. 65.9 mL, p < 0.001) than women. Diastolic indices differed by age and sex, such that women and older individuals had higher E/e′ ratio (p<0.001) (Figure 1B). No significant sex differences were observed in the prevalence of clinically defined DD grade (p = 0.768); however, it differed significantly across age groups (p = 0.039). Although older participants (age 65-83) also had significantly higher LAVI (p< 0.001) (Figure 1C), it did not differ by sex. Structural measures showed a higher LVMI in men than women (84.5 vs. 74.9 g/m^2^, p< 0.001) and older (age 65-83) than younger participants (age 41-57) (81.9 vs 76.9, p<0.001) (Figure 1D). Women tended to have higher prevalence of concentric remodeling (70.6%) and LVH (8.4%) compared to men (67.9% and 6.4%, respectively); but these differences were not statistically significant. However, the difference of LVH was significantly greater in older participants compared to younger participants (9.1% vs 4.2%, p=0.005). The mean value of aortic valve area and peak velocity were in the normal range for all groups, but the prevalence of aortic stenosis was borderline significantly higher in men than women (3.9% vs. 1.8% p=0.055). We also subdivided the men and women according to median age 60 years and further analyzed the echocardiographic characteristics (Table 3). Most of the echocardiographic measures in men fell withing the normal range and did not differ significantly. However, several measurements significantly differ between <60 and ≥60 years of aged older women. Structural abnormalities, including concentric remodeling (72.7%) and LVH (9.1%) were significantly higher women in ≥60 years old than in women <60 years of age. Similarly, the E/e’ ratio was significantly higher in women ≥60 years old than women <60 years of age (10.1 vs 8.9, p<0.001) (Figure 2A). Although LVEF differed significantly between men and women, it remained largely preserved across all age groups and both sexes (Figure 2B).

**Figure 1:**
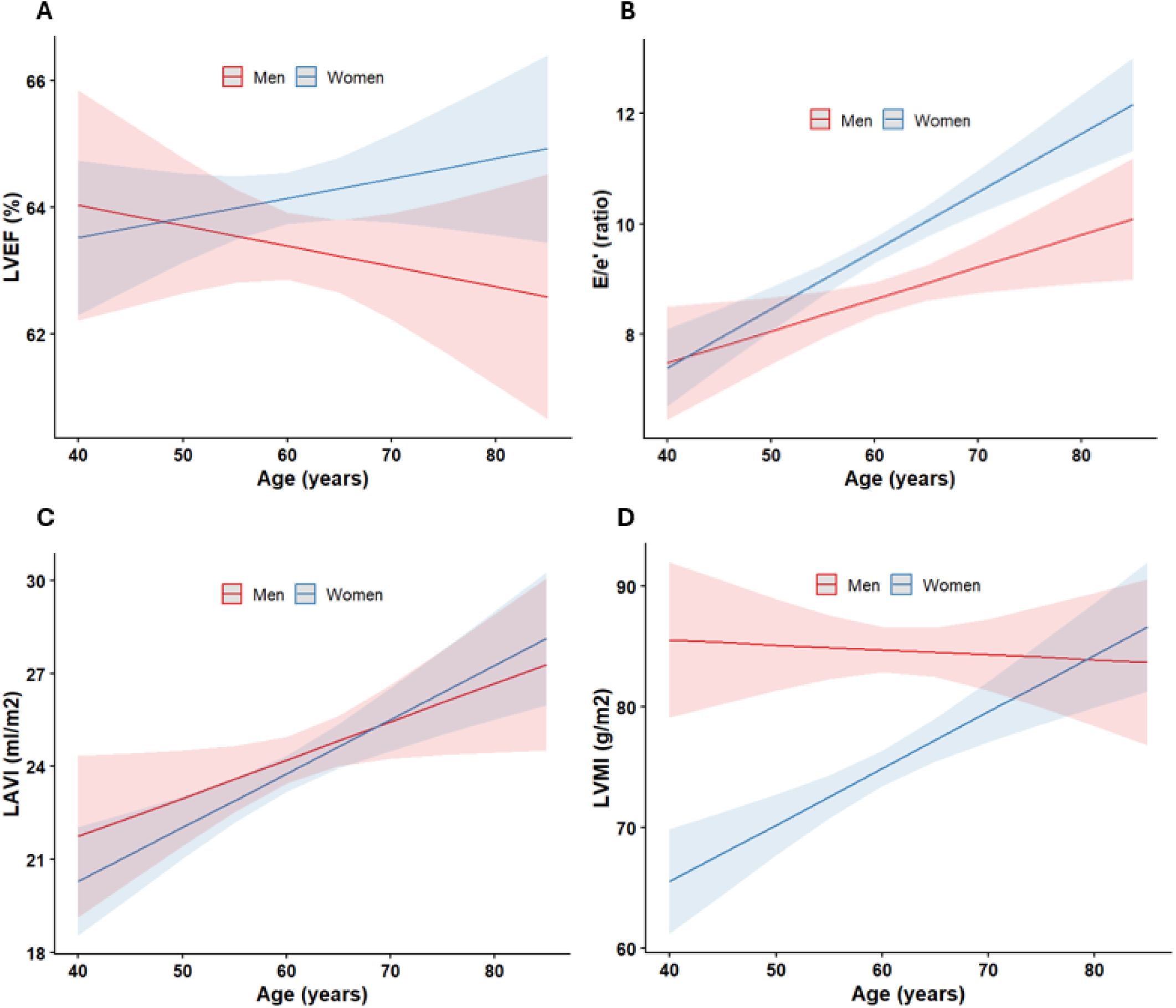
Age and sex specific Left Ventricular Ejection Fraction (LVEF), E/e’ ratio, Left Atrium Volume Index (LAVI) and Left Ventricular Mass Index (LVMI) in THS. (A) LVEF was higher and increased with age in women than men. (B) Diastolic indices E/e’ ratio also increased with age for both men and women; but it increased higher in women than men. (C) LAVI increase with age for both sex groups. (D) LVMI increased with age in women than men.

**Figure 2:**
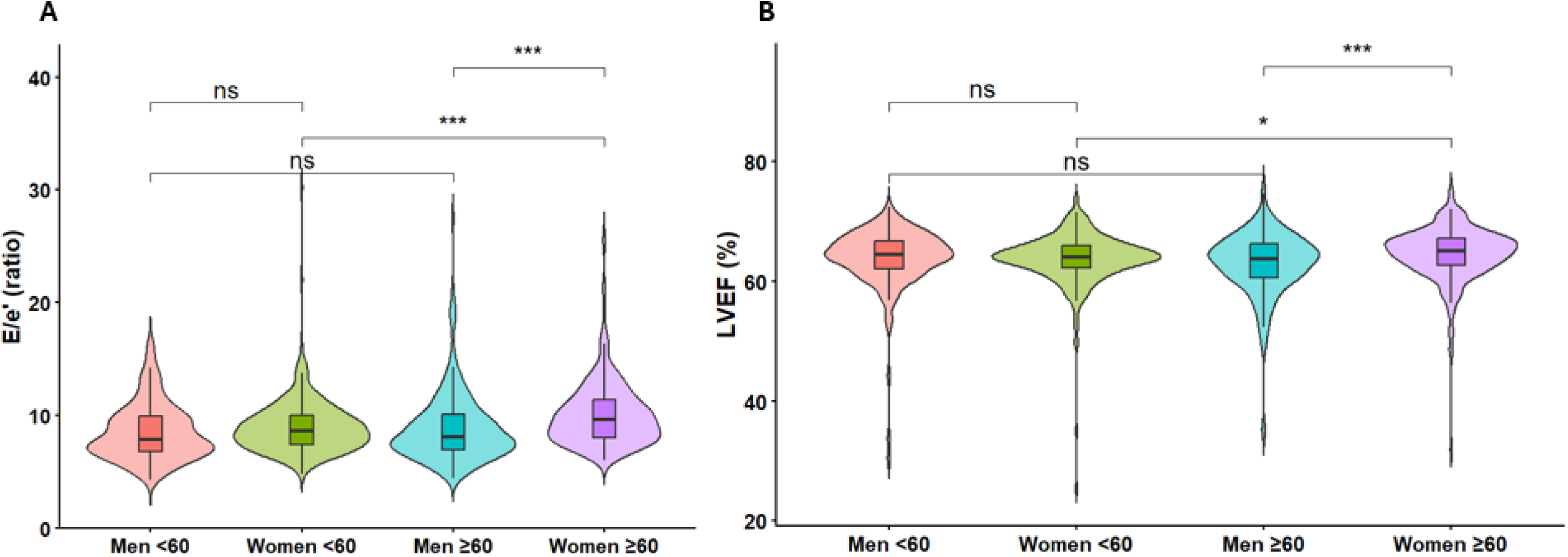
Association of E/e’ ratio and Left Ventricular Ejection Fraction (LVEF) between sex and age group (<60 and ≥60). A) E/e’ ratio was significantly higher in women ≥60 years old than women <60 years of age and men ≥60 years old. B) LVEF was also significantly higher in women ≥60 years old than women <60 years of age and men ≥60 years old.

**Table 3:** Comparisons of cardiac structure and function measures between younger and older men and women in the THS.

| Echocardiographic measures<br>[mean (SD) or %, as appropriate] | Men |  |  | Women |  |  | Sex difference p-value<sup>1</sup> |
|---|---|---|---|---|---|---|---|
|  | Age <60<br>[N=151] | Age ≥60<br>[N=206] | p-value | Age <60<br>[N=294] | Age ≥60<br>[N=273] | p-value |  |
| ***LV structure*** |  |  |  |  |  |  |  |
| LV internal diameter diastole (mm) | 44.6 (4.9) | 44.2 (5.0) | 0.480 | 41.9 (4.7) | 41.3 (4.7) | 0.093 | <0.001 |
| LV internal diameter systole (mm) | 28.7 (4.3) | 28.8 (4.3) | 0.811 | 26.9 (3.8) | 26.2 (3.6) | 0.034 | <0.001 |
| LVEDV Index to BSA (ml/m<sup>2</sup>) | 38.4 (8.9) | 38.8 (8.4) | 0.666 | 36.3 (7.4) | 34.2 (6.6) | 0.004 | <0.001 |
| LVEDV (ml) | 76.9 (18.6) | 75.4 (15.9) | 0.406 | 69.5 (15.6) | 63.5 (13.2) | <0.001 | <0.001 |
| LVESV Index to BSA (ml/m<sup>2</sup>) | 14.1 (5.2) | 14.5 (4.5) | 0.439 | 13.3 (4.4) | 12.1 (3.2) | 0.001 | <0.001 |
| LVESV (ml) | 28.3 (11.6) | 28.1 (8.4) | 0.847 | 25.4 (8.9) | 22.5 (6.3) | <0.001 | <0.001 |
| LV septal wall thickness diastole (mm) | 11.1 (1.4) | 10.9 (1.4) | 0.094 | 10.3 (1.3) | 10.7 (1.4) | <0.001 | <0.001 |
| LV posterior wall thickness diastole (mm) | 10.5 (1.4) | 10.4 (1.4) | 0.476 | 9.8 (1.2) | 10.2 (1.3) | <0.001 | <0.001 |
| LV septal wall thickness systole (mm) | 14.9 (1.7) | 14.6 (1.7) | 0.097 | 13.5 (1.3) | 13.8 (1.6) | 0.004 | <0.001 |
| LV posterior wall thickness systole (mm) | 14.8 (1.6) | 14.5 (1.6) | 0.067 | 13.5 (1.3) | 13.8 (1.6) | 0.004 | <0.001 |
| LV relative wall thickness (%) | 48.0 (9.2) | 47.9 (8.9) | 0.936 | 47.2 (7.8) | 50.1 (9.2) | <0.001 | 0.282 |
| LV mass (g) | 171.0 (40.6) | 165.0 (43.0) | 0.194 | 139.0 (32.7) | 145.0 (35.6) | 0.052 | <0.001 |
| LVMI to BSA (g/m<sup>2</sup>) | 84.8 (18.0) | 84.5 (20.5) | 0.872 | 72.3 (13.7) | 77.8 (17.9) | <0.001 | <0.001 |
| Normal geometry (%), referent | 25.8 | 25.9 | -- | 25.5 | 15.9 | -- | -- |
| Concentric remodeling (%) | 67.5 | 67.8 | 0.869 | 68.6 | 72.7 | 0.015 | 0.172 |
| Concentric LVH (%) | 5.3 | 5.9 | 0.786 | 4.1 | 9.1 | 0.002 | 0.284 |
| Eccentric LVH (%) | 1.3 | 0.5 | 0.439 | 1.7 | 2.3 | 0.239 | 0.117 |
| ***LV function*** |  |  |  |  |  |  |  |
| LV outflow VTI (cm) | 22.1 (5.5) | 21.5 (4.4) | 0.278 | 22.8 (4.2) | 23.5 (4.6) | 0.064 | <0.001 |
| Stroke volume (ml) | 71.6 (18.6) | 69.8 (15.7) | 0.348 | 66.0 (14.3) | 65.8 (13.8) | 0.811 | <0.001 |
| Stroke volume index to BSA (ml/m<sup>2</sup>) | 35.7 (9.8) | 35.9 (8.2) | 0.891 | 34.6 (7.2) | 35.5 (7.2) | 0.144 | 0.147 |
| Ejection fraction (%) | 63.8 (5.4) | 63.0 (5.3) | 0.149 | 63.7 (4.5) | 64.6 (4.5) | 0.023 | 0.021 |
| Ejection fraction categories |  |  |  |  |  |  |  |
| ≥60% | 88.1 | 79.4 |  | 87.7 | 89.7 |  |  |
| 50%-59.9% | 9.9 | 18.6 | 0.147 | 10.9 | 9.2 | 0.861 | 0.109 |
| 40%-49.9% | 0.7 | 1.0 |  | 0.7 | 0.7 |  |  |
| <40% | 1.3 | 1.0 |  | 0.7 | 0.4 |  |  |
| LV TDI s' lateral (m/s) | 0.09 (0.02) | 0.09 (0.02) | 0.585 | 0.08 (0.02) | 0.07 (0.02) | 0.004 | <0.001 |
| GLS (%) | -21.7 (2.8) | -21.6 (2.7) | 0.703 | -22.5 (2.4) | -22.3 (2.8) | 0.549 | <0.001 |
| GLS categories |  |  |  |  |  |  |  |
| Normal (≤-18%) | 94.5 | 91.9 |  | 97.6 | 94.1 |  |  |
| Mildly impaired (-16%-17.9%) | 4.7 | 4.9 | 0.353 | 2.4 | 4.6 | 0.074 | 0.080 |
| Severely impaired (>16%) | 0.8 | 3.2 |  | 0.0 | 1.3 |  |  |
| LV TDI e' lateral (m/s) | 0.09 (0.03) | 0.08 (0.03) | 0.386 | 0.10 (0.03) | 0.09 (0.02) | <0.001 | 0.261 |
| LV TDI e' septal (m/s) | 0.07 (0.02) | 0.07 (0.02) | 0.618 | 0.08 (0.02) | 0.07 (0.02) | <0.001 | <0.001 |
| LV TDI A' lateral (m/s) | 0.10 (0.03) | 0.10 (0.03) | 0.804 | 0.10 (0.03) | 0.10 (0.02) | 0.141 | 0.013 |
| LV TDI A' septal (m/s) | 0.10 (0.02) | 0.10 (0.02) | 0.381 | 0.09 (0.02) | 0.10 (0.02) | 0.515 |  |
| Diastolic dysfunction (%) |  |  |  |  |  |  |  |
| None | 88.5 | 83.1 |  | 87.8 | 87.0 |  |  |
| Grade 1 | 10.1 | 13.9 | 0.462 | 11.5 | 10.3 | 0.236 | 0.768 |
| Grade 2 | 1.4 | 2.5 |  | 0.3 | 2.3 |  |  |
| Grade 3 | 0 | 0.5 |  | 0.3 | 0.4 |  |  |
| ***RV structure*** |  |  |  |  |  |  |  |
| RV diameter at base (mm) | 35.4 (4.3) | 35.0 (4.7) | 0.447 | 33.7 (3.9) | 33.7 (4.1) | 0.949 | <0.001 |
| RV end diastolic area (cm<sup>2</sup>) | 14.7 (2.8) | 14.4 (3.3) | 0.335 | 13.7 (2.9) | 13.2 (3.2) | 0.110 | <0.001 |
| RV end systolic area (cm<sup>2</sup>) | 8.1 (1.6) | 8.0 (2.4) | 0.603 | 7.5 (1.8) | 7.2 (1.9) | 0.163 | <0.001 |
| ***RV function*** |  |  |  |  |  |  |  |
| RV fractional area change (%) | 44.7 (3.7) | 44.5 (5.1) | 0.783 | 45.3 (5.2) | 45.3 (5.0) | 0.978 | 0.034 |
| TAPSE (mm) | 21.8 (3.3) | 22.1 (3.2) | 0.379 | 21.8 (2.7) | 22.1 (2.7) | 0.348 | 0.868 |
| S' (m/s) | 0.14 (0.03) | 0.13 (0.02) | 0.118 | 0.13 (0.02) | 0.14 (0.02) | 0.191 | 0.975 |
| RVOT VTI | 16.0 (3.6) | 15.4 (3.1) | 0.149 | 16.9 (3.1) | 16.9 (3.6) | 0.819 | <0.001 |
| ***LA structure and function*** |  |  |  |  |  |  |  |
| LA diameter (mm) | 32.9 (4.5) | 32.5 (4.2) | 0.490 | 31.9 (3.9) | 32.3 (3.9) | 0.237 | 0.033 |
| LA volume (ml) | 46.8 (13.5) | 49.1 (15.8) | 0.139 | 43.4 (13.0) | 46.5 (12.7) | 0.005 | 0.001 |
| LAVI to BSA (ml/m<sup>2</sup>) | 23.3 (6.4) | 25.3 (8.1) | 0.012 | 22.7 (6.6) | 24.9 (6.6) | <0.001 | 0.191 |
| ***Mitral valve*** |  |  |  |  |  |  |  |
| Peak E velocity (m/s) | 0.6 (0.2) | 0.7 (0.2) | 0.831 | 0.8 (0.2) | 0.8 (0.2) | 0.988 | <0.001 |
| Peak A velocity (m/s) | 0.8 (0.2) | 0.8 (0.2) | 0.119 | 0.8 (0.2) | 0.9 (0.2) | <0.001 | <0.001 |
| E/A ratio | 0.9 (0.2) | 0.9 (0.3) | 0.404 | 0.9 (0.2) | 0.8 (0.2) | 0.003 | 0.013 |
| ***Aortic valve*** |  |  |  |  |  |  |  |
| Peak velocity (m/s) | 1.3 (0.4) | 1.3 (0.4) | 0.431 | 1.3 (0.2) | 1.3 (0.2) | 0.405 | 0.947 |
| Mean gradient | 17.3 (5.3) | 19.8 (5.4) | <0.001 | 19.1 (4.6) | 21.1 (5.8) | <0.001 | 0.001 |
| Valve area (cm<sup>2</sup>) | 2.6 (0.6) | 2.6 (0.6) | 0.478 | 2.4 (0.5) | 2.3 (0.4) | 0.031 | <0.001 |
| Aortic stenosis |  |  |  |  |  |  |  |
| No (%) | 96.0 | 96.6 | 0.99 | 99.0 | 98.2 | 0.644 | 0.027 |
| Yes (%) | 4.0 | 3.4 |  | 1.0 | 1.8 |  |  |
| ***Hemodynamics*** |  |  |  |  |  |  |  |
| Cardiac output (l/min) | 4.7 (1.4) | 4.5 (1.1) | 0.115 | 4.6 (1.1) | 4.5 (1.1) | 0.095 | 0.493 |
| Cardiac index to BSA (l/min/m<sup>2</sup>) | 2.4 (0.7) | 2.3 (0.6) | 0.580 | 2.4 (0.6) | 2.4 (0.6) | 0.829 | 0.032 |
| Average E/e'(ratio) | 8.5 (2.5) | 8.9 (3.2) | 0.234 | 8.9 (2.5) | 10.1 (2.9) | <0.001 | <0.001 |
| TR velocity (m/s) | 2.1 (0.3) | 2.2 (0.3) | <0.001 | 2.2 (0.3) | 2.3 (0.3) | 0.001 | 0.001 |
| PASP (mm Hg) | 22.3 (5.3) | 34.8 (5.4) | <0.001 | 24.1 (4.6) | 26.1 (5.8) | <0.001 | 0.001 |
| Pulmonary vascular resistance (WU) | 1.5 (0.4) | 1.6 (0.4) | 0.003 | 1.5 (0.3) | 1.6 (0.4) | 0.011 | 0.014 |

SD=Standard Deviation; LV=Left Ventricular; LVEDV=Left Ventricular End Diastolic Volume; BSA=Body Surface Area; LVMI= Left Ventricular Mass Index; LA=Left Atrium; LAVI= Left Atrium Volume Index; RV=Right Ventricular; TAPSE=Tricuspid Annular Plane Systolic Excursion; GLS=Global Longitudinal Strain; RVOT VTI=Right Ventricular Outflow Tract Velocity-Time Integral; TDI=Tissue Doppler Imaging; TR= Tricuspid Regurgitation; PASP= Pulmonary Artery Systolic Pressure. It was calculated using TR velocity and right atrial pressure of 5 mm Hg. <sup>1</sup>t-test for continuous outcomes and chi-square test for dichotomous outcomes were used to calculate the p-value.

**Table 4:**
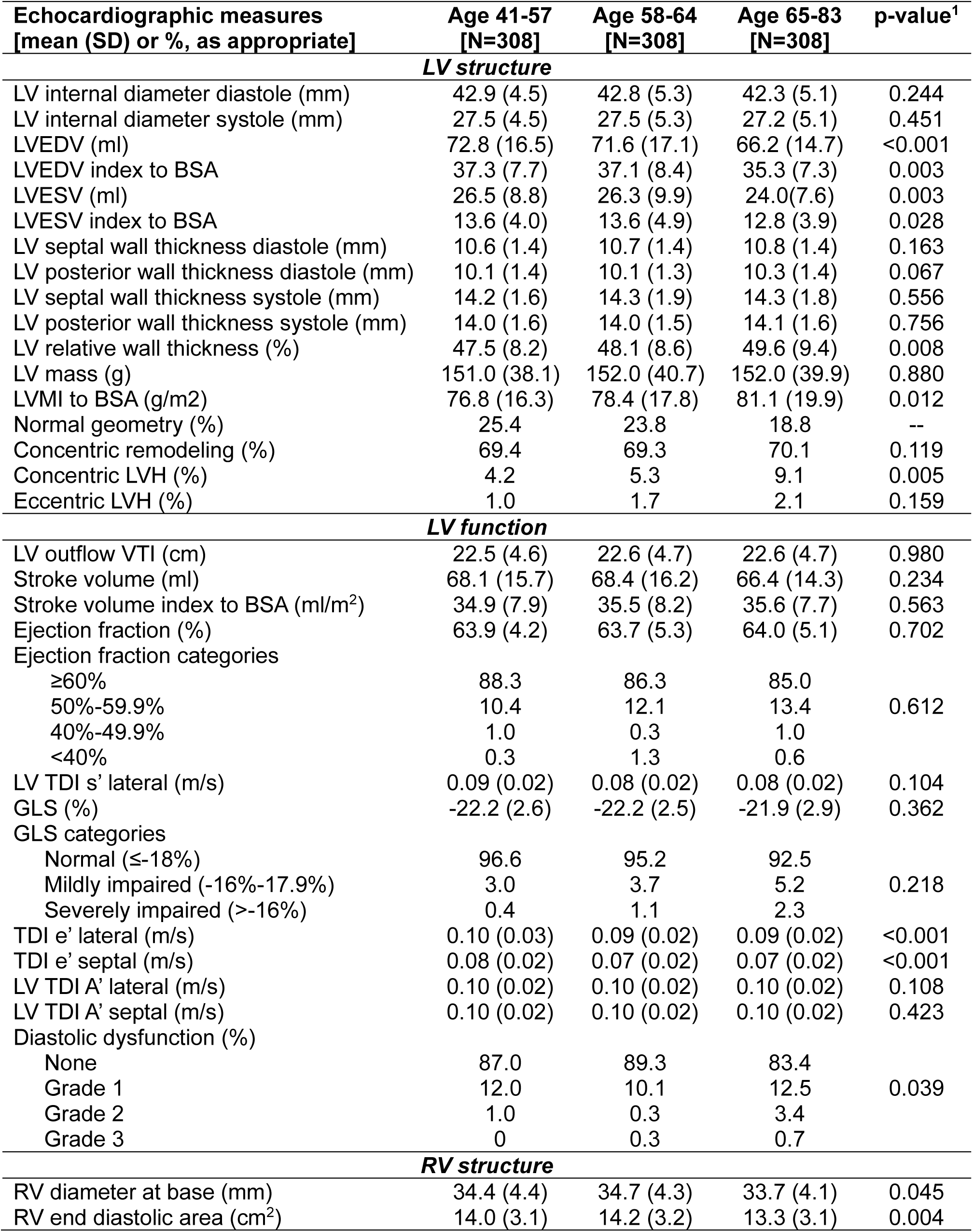
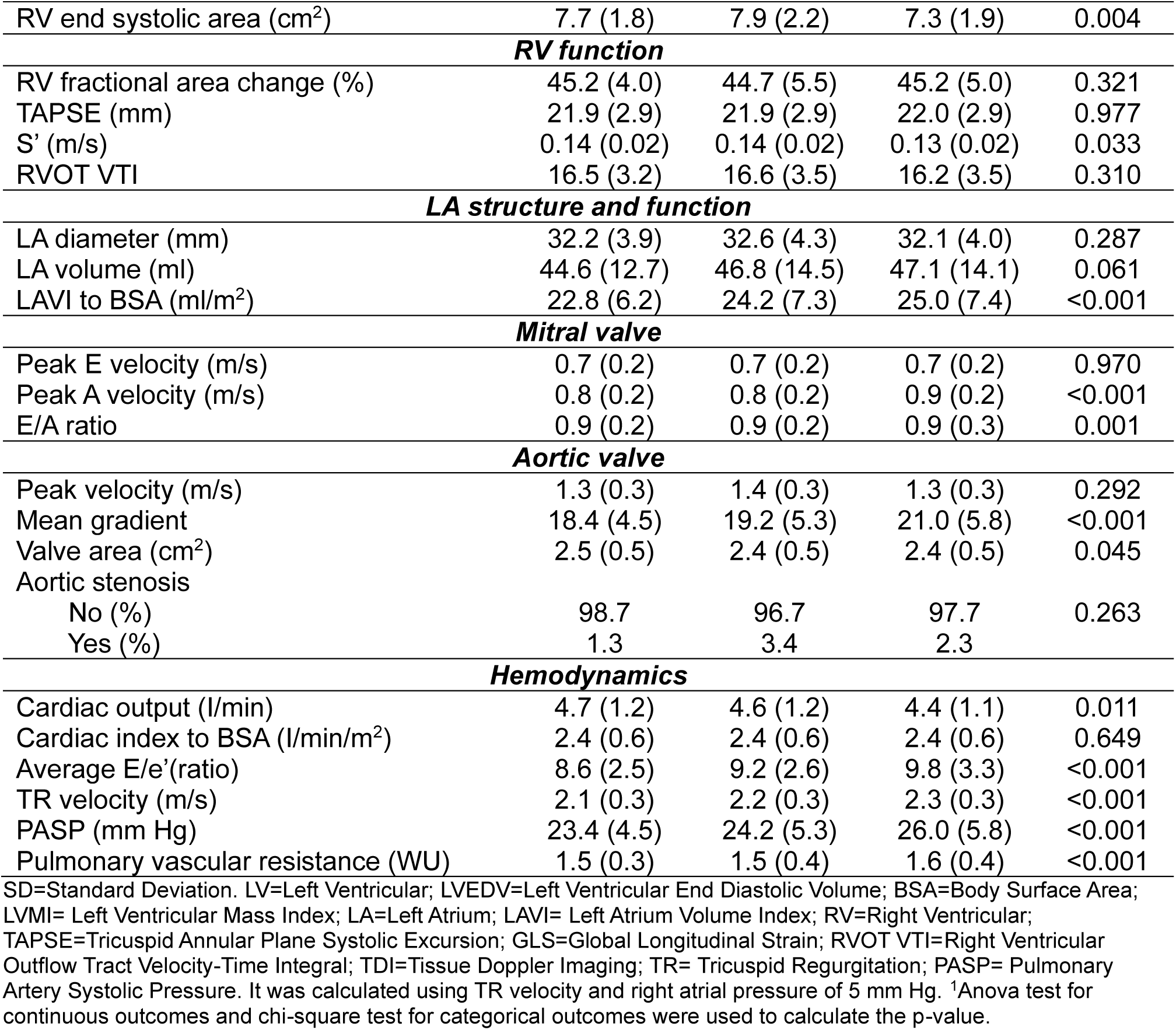
Age specific comparisons of cardiac structure and function measures in the THS.

| **Echocardiographic measures<br>[mean (SD) or %, as appropriate]** | **Age 41-57<br>[N=308]** | **Age 58-64<br>[N=308]** | **Age 65-83<br>[N=308]** | **p-value<sup>1</sup>** |
|---|---|---|---|---|
| ***LV structure*** |  |  |  |  |
| LV internal diameter diastole (mm) | 42.9 (4.5) | 42.8 (5.3) | 42.3 (5.1) | 0.244 |
| LV internal diameter systole (mm) | 27.5 (4.5) | 27.5 (5.3) | 27.2 (5.1) | 0.451 |
| LVEDV (ml) | 72.8 (16.5) | 71.6 (17.1) | 66.2 (14.7) | <0.001 |
| LVEDV index to BSA | 37.3 (7.7) | 37.1 (8.4) | 35.3 (7.3) | 0.003 |
| LVESV (ml) | 26.5 (8.8) | 26.3 (9.9) | 24.0(7.6) | 0.003 |
| LVESV index to BSA | 13.6 (4.0) | 13.6 (4.9) | 12.8 (3.9) | 0.028 |
| LV septal wall thickness diastole (mm) | 10.6 (1.4) | 10.7 (1.4) | 10.8 (1.4) | 0.163 |
| LV posterior wall thickness diastole (mm) | 10.1 (1.4) | 10.1 (1.3) | 10.3 (1.4) | 0.067 |
| LV septal wall thickness systole (mm) | 14.2 (1.6) | 14.3 (1.9) | 14.3 (1.8) | 0.556 |
| LV posterior wall thickness systole (mm) | 14.0 (1.6) | 14.0 (1.5) | 14.1 (1.6) | 0.756 |
| LV relative wall thickness (%) | 47.5 (8.2) | 48.1 (8.6) | 49.6 (9.4) | 0.008 |
| LV mass (g) | 151.0 (38.1) | 152.0 (40.7) | 152.0 (39.9) | 0.880 |
| LVMI to BSA (g/m2) | 76.8 (16.3) | 78.4 (17.8) | 81.1 (19.9) | 0.012 |
| Normal geometry (%) | 25.4 | 23.8 | 18.8 | -- |
| Concentric remodeling (%) | 69.4 | 69.3 | 70.1 | 0.119 |
| Concentric LVH (%) | 4.2 | 5.3 | 9.1 | 0.005 |
| Eccentric LVH (%) | 1.0 | 1.7 | 2.1 | 0.159 |
| ***LV function*** |  |  |  |  |
| LV outflow VTI (cm) | 22.5 (4.6) | 22.6 (4.7) | 22.6 (4.7) | 0.980 |
| Stroke volume (ml) | 68.1 (15.7) | 68.4 (16.2) | 66.4 (14.3) | 0.234 |
| Stroke volume index to BSA (ml/m<sup>2</sup>) | 34.9 (7.9) | 35.5 (8.2) | 35.6 (7.7) | 0.563 |
| Ejection fraction (%) | 63.9 (4.2) | 63.7 (5.3) | 64.0 (5.1) | 0.702 |
| Ejection fraction categories |  |  |  |  |
| ≥60% | 88.3 | 86.3 | 85.0 |  |
| 50%-59.9% | 10.4 | 12.1 | 13.4 | 0.612 |
| 40%-49.9% | 1.0 | 0.3 | 1.0 |  |
| <40% | 0.3 | 1.3 | 0.6 |  |
| LV TDI s’ lateral (m/s) | 0.09 (0.02) | 0.08 (0.02) | 0.08 (0.02) | 0.104 |
| GLS (%) | -22.2 (2.6) | -22.2 (2.5) | -21.9 (2.9) | 0.362 |
| GLS categories |  |  |  |  |
| Normal (≤-18%) | 96.6 | 95.2 | 92.5 |  |
| Mildly impaired (-16%-17.9%) | 3.0 | 3.7 | 5.2 | 0.218 |
| Severely impaired (>-16%) | 0.4 | 1.1 | 2.3 |  |
| TDI e’ lateral (m/s) | 0.10 (0.03) | 0.09 (0.02) | 0.09 (0.02) | <0.001 |
| TDI e’ septal (m/s) | 0.08 (0.02) | 0.07 (0.02) | 0.07 (0.02) | <0.001 |
| LV TDI A’ lateral (m/s) | 0.10 (0.02) | 0.10 (0.02) | 0.10 (0.02) | 0.108 |
| LV TDI A’ septal (m/s) | 0.10 (0.02) | 0.10 (0.02) | 0.10 (0.02) | 0.423 |
| Diastolic dysfunction (%) |  |  |  |  |
| None | 87.0 | 89.3 | 83.4 |  |
| Grade 1 | 12.0 | 10.1 | 12.5 | 0.039 |
| Grade 2 | 1.0 | 0.3 | 3.4 |  |
| Grade 3 | 0 | 0.3 | 0.7 |  |
| ***RV structure*** |  |  |  |  |
| RV diameter at base (mm) | 34.4 (4.4) | 34.7 (4.3) | 33.7 (4.1) | 0.045 |
| RV end diastolic area (cm<sup>2</sup>) | 14.0 (3.1) | 14.2 (3.2) | 13.3 (3.1) | 0.004 |
| RV end systolic area (cm<sup>2</sup>) | 7.7 (1.8) | 7.9 (2.2) | 7.3 (1.9) | 0.004 |
| ***RV function*** |  |  |  |  |
| RV fractional area change (%) | 45.2 (4.0) | 44.7 (5.5) | 45.2 (5.0) | 0.321 |
| TAPSE (mm) | 21.9 (2.9) | 21.9 (2.9) | 22.0 (2.9) | 0.977 |
| S’ (m/s) | 0.14 (0.02) | 0.14 (0.02) | 0.13 (0.02) | 0.033 |
| RVOT VTI | 16.5 (3.2) | 16.6 (3.5) | 16.2 (3.5) | 0.310 |
| ***LA structure and function*** |  |  |  |  |
| LA diameter (mm) | 32.2 (3.9) | 32.6 (4.3) | 32.1 (4.0) | 0.287 |
| LA volume (ml) | 44.6 (12.7) | 46.8 (14.5) | 47.1 (14.1) | 0.061 |
| LAVI to BSA (ml/m<sup>2</sup>) | 22.8 (6.2) | 24.2 (7.3) | 25.0 (7.4) | <0.001 |
| ***Mitral valve*** |  |  |  |  |
| Peak E velocity (m/s) | 0.7 (0.2) | 0.7 (0.2) | 0.7 (0.2) | 0.970 |
| Peak A velocity (m/s) | 0.8 (0.2) | 0.8 (0.2) | 0.9 (0.2) | <0.001 |
| E/A ratio | 0.9 (0.2) | 0.9 (0.2) | 0.9 (0.3) | 0.001 |
| ***Aortic valve*** |  |  |  |  |
| Peak velocity (m/s) | 1.3 (0.3) | 1.4 (0.3) | 1.3 (0.3) | 0.292 |
| Mean gradient | 18.4 (4.5) | 19.2 (5.3) | 21.0 (5.8) | <0.001 |
| Valve area (cm<sup>2</sup>) | 2.5 (0.5) | 2.4 (0.5) | 2.4 (0.5) | 0.045 |
| Aortic stenosis |  |  |  |  |
| No (%) | 98.7 | 96.7 | 97.7 | 0.263 |
| Yes (%) | 1.3 | 3.4 | 2.3 |  |
| ***Hemodynamics*** |  |  |  |  |
| Cardiac output (I/min) | 4.7 (1.2) | 4.6 (1.2) | 4.4 (1.1) | 0.011 |
| Cardiac index to BSA (I/min/m<sup>2</sup>) | 2.4 (0.6) | 2.4 (0.6) | 2.4 (0.6) | 0.649 |
| Average E/e’(ratio) | 8.6 (2.5) | 9.2 (2.6) | 9.8 (3.3) | <0.001 |
| TR velocity (m/s) | 2.1 (0.3) | 2.2 (0.3) | 2.3 (0.3) | <0.001 |
| PASP (mm Hg) | 23.4 (4.5) | 24.2 (5.3) | 26.0 (5.8) | <0.001 |
| Pulmonary vascular resistance (WU) | 1.5 (0.3) | 1.5 (0.4) | 1.6 (0.4) | <0.001 |

SD=Standard Deviation. LV=Left Ventricular; LVEDV=Left Ventricular End Diastolic Volume; BSA=Body Surface Area; LVMI= Left Ventricular Mass Index; LA=Left Atrium; LAVI= Left Atrium Volume Index; RV=Right Ventricular; TAPSE=Tricuspid Annular Plane Systolic Excursion; GLS=Global Longitudinal Strain; RVOT VTI=Right Ventricular Outflow Tract Velocity-Time Integral; TDI=Tissue Doppler Imaging; TR= Tricuspid Regurgitation; PASP= Pulmonary Artery Systolic Pressure. It was calculated using TR velocity and right atrial pressure of 5 mm Hg. <sup>1</sup>Anova test for continuous outcomes and chi-square test for categorical outcomes were used to calculate the p-value.

Echocardiographic characteristics were also compared among HF stages in Table 5. Overall, 68.7%, 19.0%, and 0.8% of participants were classified as having Stage A, Stage B, and Stage C heart failure, respectively. In addition, heart failure stages differed significantly between men and women (p = 0.002). Women had a higher prevalence of Stage A (71.6%% vs. 64.1%) and Stage B (19.4% vs. 18.5%) heart failure than men. In contrast, Stage C heart failure was more prevalent among men than women (1.1% vs. 0.5%). LV mass and LVMI were significantly higher in stage C (LV mass=218.0 g, and LVMI=114.0 g/m^2^). Stroke volume was significantly lower (57.4 mL, p=0.034) in HF stage C. Consistent with this finding, LVEF was also substantially lower (32.4%) in stage C. Moreover, there was significant difference in the E/e’ ratio among the HF stages and the people from HF stage C showed higher E/e’ ratio (14.4) compared to other stages.

**Table 5:** Heart failure stages and cardiac structure and function measures in the THS.

| **Echocardiographic measures [mean (SD) or %, as appropriate]** | **Stage 0 [N=107, 11.6%]** | **Stage A [N=634, 68.6%]** | **Stage B [N=176, 19.0%]** | **Stage C [N=7, 0.8%]** | **p-value<sup>1</sup>** |
|---|---|---|---|---|---|
| ***LV structure*** |  |  |  |  |  |
| LV internal diameter diastole (mm) | 42.0 (4.5) | 42.1 (4.7) | 44.8 (5.1) | 51.4 (12.6) | <0.001 |
| LV internal diameter systole (mm) | 26.9 (3.4) | 26.9 (3.4) | 29.1 (4.3) | 41.4 (14.3) | <0.001 |
| LVEDV (ml) | 69.9 (14.4) | 68.7 (14.9) | 74.7 (16.7) | 108.0 (59.0) | <0.001 |
| LVEDV index to BSA | 37.9 (6.9) | 35.6 (7.1) | 38.5 (8.1) | 55.3 (27.0) | 0.001 |
| LVESV (ml) | 25.3 (6.4) | 24.6 (6.5) | 27.8 (8.5) | 68.6 (50.0) | <0.001 |
| LVESV index to BSA | 13.8 (3.2) | 12.8 (3.2) | 14.3 (4.2) | 34.2 (24.0) | <0.001 |
| LV septal wall thickness diastole (mm) | 9.8 (1.2) | 10.7 (1.3) | 11.4 (1.6) | 9.9 (2.6) | <0.001 |
| LV posterior wall thickness diastole (mm) | 9.4 (1.1) | 10.1 (1.3) | 10.9 (1.4) | 11.2 (2.9) | <0.001 |
| LV septal wall thickness systole (mm) | 13.4 (1.1) | 14.2 (1.5) | 14.8 (1.9) | 18.2 (9.6) | <0.001 |
| LV posterior wall thickness systole (mm) | 13.2 (1.4) | 14.0 (1.4) | 14.6 (1.9) | 13.8 (2.8) | <0.001 |
| LV relative wall thickness (%) | 45.1 (7.6) | 48.8 (8.9) | 49.0 (7.8) | 46.8 (23.1) | 0.005 |
| LV mass (g) | 131.0 (27.1) | 147.0 (32.0) | 180.0 (49.9) | 218.0 (97.4) | <0.001 |
| LVMI to BSA (g/m2) | 71.3 (13.5) | 75.8 (14.0) | 92.4 (23.5) | 114.0 (47.6) | <0.001 |
| Normal geometry (%), referent | 36.8 | 23.2 | 12.6 | 33.3 | -- |
| Concentric remodeling (%) | 63.2 | 66.8 | 48.9 | 16.6 | 0.031 |
| Concentric LVH (%) | 0 | 0 | 31.0 | 33.3 | <0.001 |
| Eccentric LVH (%) | 0 | 0 | 7.5 | 16.6 | <0.001 |
| ***LV function*** |  |  |  |  |  |
| LV outflow VTI (cm) | 21.9 (4.2) | 22.5 (4.6) | 23.3 (5.1) | 18.1 (3.4) | 0.086 |
| Stroke volume (ml) | 66.5 (13.1) | 67.1 (15.0) | 71.0 (17.7) | 57.4 (13.2) | 0.034 |
| Stroke volume index to BSA (ml/m<sup>2</sup>) | 36.3 (6.9) | 34.9 (7.6) | 36.7 (9.3) | 30.6 (8.4) | 0.682 |
| Ejection fraction (%) | 63.9 (3.5) | 64.4 (3.6) | 63.2 (5.3) | 32.4 (3.6) | <0.001 |
| Ejection fraction categories |  |  |  |  |  |
| ≥60% | 89.6 | 89.7 | 76.7 | 0.0 |  |
| 50%-59.9% | 10.4 | 10.3 | 19.3 | 0.0 | <0.001 |
| 40%-49.9% | 0.0 | 0.0 | 4.0 | 0.0 |  |
| <40% | 0.0 | 0.0 | 0.0 | 100 |  |
| LV TDI s’ lateral (m/s) | 0.09 (0.02) | 0.08 (0.02) | 0.08 (0.02) | 0.06 (0.02) | <0.001 |
| GLS (%) | -22.2 (2.4) | -22.2 (2.5) | -21.9 (3.2) | -17.2 (3.9) | 0.059 |
| GLS categories |  |  |  |  |  |
| Normal (≤-18%) | 96.9 | 96.0 | 89.9 | 60.0 |  |
| Mildly impaired (-16%-17.9%) | 2.1 | 3.8 | 6.1 | 0.0 | <0.001 |
| Severely impaired (>16%) | 1.0 | 0.2 | 4.0 | 40.0 |  |
| LV TDI e’ lateral (m/s) | 0.1 (0.02) | 0.09 (0.02) | 0.09 (0.03) | 0.07 (0.03) | 0.001 |
| LV TDI e’ septal (m/s) | 0.09 (0.02) | 0.07 (0.02) | 0.07 (0.02) | 0.05 (0.02) | <0.001 |
| LV TDI A’ lateral (m/s) | 0.10 (0.02) | 0.10 (0.02) | 0.09 (0.02) | 0.08 (0.03) | 0.001 |
| LV TDI A’ septal (m/s) | 0.10 (0.02) | 0.10 (0.02) | 0.09 (0.02) | 0.08 (0.02) | <0.001 |
| Diastolic dysfunction (%) |  |  |  |  | <0.001 |
| None | 100.0 | 100.0 | 31.8 | 50.0 |  |
| Grade 1 | 0 | 0 | 59.5 | 0 |  |
| Grade 2 | 0 | 0 | 7.5 | 25.0 |  |
| Grade 3 | 0 | 0 | 1.2 | 25.0 |  |
| ***RV structure*** |  |  |  |  |  |
| RV diameter at base (mm) | 34.8 (4.5) | 33.9 (4.0) | 35.0 (4.9) | 34.9 (3.7) | 0.412 |
| RV end diastolic area (cm<sup>2</sup>) | 14.1 (3.2) | 13.6 (2.3) | 14.5 (3.7) | 15.7 (3.8) | 0.067 |
| RV end systolic area (cm<sup>2</sup>) | 7.8 (2.0) | 7.5 (1.7) | 8.1 (2.7) | 7.1 (4.3) | 0.153 |
| ***RV function*** |  |  |  |  |  |
| RV fractional area change (%) | 45.2 (4.4) | 45.0 (4.2) | 44.6 (5.1) | 54.2 (24.6) | 0.602 |
| TAPSE (mm) | 22.0 (3.1) | 21.9 (2.9) | 22.2 (2.9) | 21.5 (3.8) | 0.609 |
| S’ (m/s) | 0.13 (0.02) | 0.13 (0.02) | 0.14 (0.02) | 0.13 (0.03) | 0.082 |
| RVOT VTI | 16.0 (3.2) | 16.5 (3.4) | 16.7 (3.3) | 13.4 (7.3) | 0.283 |
| ***LA structure and function*** |  |  |  |  |  |
| LA diameter (mm) | 30.2 (3.2) | 32.1 (3.8) | 34.2 (4.4) | 40.6 (9.1) | <0.001 |
| LA volume (ml) | 42.2 (10.8) | 43.9 (10.5) | 55.6 (18.0) | 72.4 (42.4) | <0.001 |
| LAVI to BSA (ml/m<sup>2</sup>) | 22.9 (5.3) | 22.7 (5.1) | 28.8 (9.6) | 37.9 (21.0) | <0.001 |
| ***Mitral valve*** |  |  |  |  |  |
| Peak E velocity (m/s) | 0.7 (0.2) | 0.7 (0.2) | 0.8 (0.2) | 0.9 (0.3) | <0.001 |
| Peak A velocity (m/s) | 0.7 (0.2) | 0.9 (0.2) | 0.8 (0.2) | 0.9 (0.3) | 0.005 |
| E/A ratio | 1.0 (0.3) | 0.8 (0.2) | 1.0 (0.3) | 1.1 (0.8) | 0.339 |
| ***Aortic valve*** |  |  |  |  |  |
| Peak velocity (m/s) | 1.2 (0.2) | 1.3 (0.3) | 1.4 (0.3) | 1.1 (0.2) | 0.003 |
| Mean gradient | 18.0 (4.4) | 19.2 (4.9) | 22.3 (6.9) | 21.2 (8.6) | <0.001 |
| Valve area (cm<sup>2</sup>) | 2.6 (0.5) | 2.4 (0.5) | 2.4 (0.6) | 2.6 (0.5) | 0.117 |
| Aortic stenosis |  |  |  |  |  |
| No (%) | 99.1 | 97.9 | 96.0 | 100.0 | 0.336 |
| Yes (%) | 0.9 | 2.1 | 4.0 | 0.0 |  |
| ***Hemodynamics*** |  |  |  |  |  |
| Cardiac output (l/min) | 4.2 (0.8) | 4.6 (1.2) | 4.8 (1.3) | 4.4(0.9) | <0.001 |
| Cardiac index to BSA (l/min/m<sup>2</sup>) | 2.3 (0.5) | 2.4 (0.6) | 2.5 (0.7) | 2.3 (0.7) | 0.012 |
| Average E/e’(ratio) | 7.9 (1.8) | 9.1 (2.5) | 10.4 (3.8) | 14.4 (1.1) | <0.001 |
| TR velocity (m/s) | 2.1 (0.3) | 2.2 (0.3) | 2.3 (0.4) | 2.3 (0.4) | <0.001 |
| PASP (mm Hg) | 23.0 (4.4) | 24.2 (4.9) | 27.3 (6.9) | 26.2 (8.6) | <0.001 |
| Pulmonary vascular resistance (WU) | 1.5 (0.3) | 1.5 (0.3) | 1.6 (0.4) | 2.1 (1.1) | 0.012 |

SD=Standard Deviation. LV=Left Ventricular; LVEDV=Left Ventricular End Diastolic Volume; BSA=Body Surface Area; LVMI= Left Ventricular Mass Index; LA=Left Atrium; LAVI= Left Atrium Volume Index; RV=Right Ventricular; TAPSE=Tricuspid Annular Plane Systolic Excursion; GLS=Global Longitudinal Strain; RVOT VTI=Right Ventricular Outflow Tract Velocity-Time Integral; TDI=Tissue Doppler Imaging; TR= Tricuspid Regurgitation; PASP= Pulmonary Artery Systolic Pressure. It was calculated using TR velocity and right atrial pressure of 5 mm Hg.<sup>1</sup>Anova test for continuous outcomes and chi-square test for dichotomous outcomes were used to calculate the p-value.

## Discussion

People with African genetic ancestry are at high risk of hypertension and abnormal cardiac structure and function ^22^. However, little is known about cardiac structure and function among the African diaspora in Tobago. Therefore, this cohort study provides the prospective standardized assessment of the epidemiology of cardiac structure and function among middle-aged to elderly African Caribbeans in the Tobago Health Study.

Hypertension is highly prevalent in the African diaspora population and one of the strongest predictors of echocardiographic abnormalities ^23^. Evidence suggests that persons with African genetic ancestry are more likely to carry gene variants associated with renal sodium retention, making blood pressure rise more strongly in response to sodium intake than those without the variants ^24^. Some studies have also showed that gene polymorphism related to ion channels, stress response, and inflammation contributed to blood pressure susceptibility and subsequent hypertension ^25,26^. Additionally, in multi-cultural communities, differences in socioeconomic status and healthcare access are also known strong determinants of hypertension burden ^27^. Chronic hypertension leads to myocardial hypertrophy, increased left ventricular mass, impaired ventricular relaxation, and progressive structural and functional remodeling of the heart ^28^. However, the impact of the high burden of hypertension on cardiac structure and function, has not been previously characterized in this population segment in the Caribbean.

We found the overall prevalence of LVH was 7.6% in THS, with greater frequency in relation to older age, rather than by sex. The prevalence of LVH we observed in THS was lower than that reported in African American population in the Jackson Heart Study (JHS) (13.9%) and the Atherosclerosis Risk in Communities (ARIC) study (12.0%) ^29,30^, as well as Caribbean population in Haiti (39.0%) ^31^. This might be due to variations in study populations, including differences in age distribution and cardiovascular risk factor burden. Notably, in the Haitian study, TTE was performed only among individuals with hypertension or CVD symptoms, which potentially contributed to the higher LVH prevalence ^31^. Although LVH was less common in THS than these other cohorts, abnormal LV geometry was very common in THS with concentric remodeling present in nearly 70%. This is likely related to the high burden of hypertension and other cardiometabolic comorbidities in this African Caribbean population ^32^ as concentric remodeling is one of the early cardiac responses to chronic hypertension ^33^.

In THS, we also found variation according to age and sex in left sided filling pressures, as measured by echocardiographic E/e’. Older individuals (age 65-83) had higher E/e’ ratio compared to younger individuals (age 41-57). This is congruent with findings from other studies, as aging is associated with LV structural changes, high collagen deposition, and myocardial fibrosis, which impairs the LV’s ability to relax efficiently during diastole and increase ventricular stiffness and filling pressure over time ^34,35^. Moreover, we found higher E/e’ ratio in women compared with men, which may be due to differences in cardiac structure, hormonal influences or generally greater susceptibility to diastolic dysfunction in women ^36^.

In this community-based cohort, the prevalences of self-reported history of myocardial infarction, stroke, atrial fibrillation, or heart failure, were low, at less than 3% for each. This may account for our finding that LVEF <50% was uncommon in the THS; which is congruent with other population-based cohort studies ^37,38^. Additionally, the mean LVEF of 63.8% in THS is consistent with findings from the JHS, ARIC and the Multi-Ethnic Study of Atherosclerosis cohorts, all of which reported largely preserved LVEF, with mean values exceeding 60% ^29,30,39^. We also found LV volumes and stroke volume were not only comparable to normal ranges reported from other populations, but also with the expected lower values in women, compared with men ^36,40^.

In THS, we found the prevalence of LVEF<40% and Stage C HF was 0.8%, while 20% of participants met Stage B, and another 69% met stage A HF criterion. Less than 12% of adults in the population-based THS classified as Stage 0. Women were more likely to have Stage A and Stage B HF, whereas men were more frequently classified as both Stage 0 and Stage C. This pattern may reflect the higher prevalence of hypertension, obesity, and diastolic dysfunction among women ^41^. On the other hand, men had a higher burden of past ischemic heart disease and myocardial infarction, which may underlie their greater prevalence of Stage C HF ^42^. Of note, given the population-based nature of this work, the presence of potential cardiac symptoms was not routinely captured, which limits our ability to report on symptomatic HF prevalence.

### Strengths

This is the first population-based cohort study that has investigated echocardiographic assessment of cardiac structure and function among African-Caribbean population in Tobago. The findings of this study will provide a baseline foundation for further longitudinal studies to establish stronger causal relationships and may also help to influence public health priorities. In addition, this study included a large number of participants with a representative part of men and women and broad age ranges, which allowed us to explore the description of cardiac structure and function between sexes and across ages. We used a rigorous approach to capturing echocardiography measures including a centralized reading lab in the U.S. using standardized image acquisition and image analysis methods similar to those used in other U.S. based cohort studies of cardiac structure and function, which may aid in future cross-cohort harmonization and analyses.

### Limitations

This study used self-reported information of medical diagnoses (e.g. stroke, myocardial infarction, diabetes, atrial fibrillation etc.), which could introduce recall bias related to history of disease. In this THS, men had a higher prevalence of Stage 0 and Stage C HF than women. The greater proportion of men classified as Stage 0 may also reflect underreporting of cardiovascular comorbidities among men compared with women. In addition, biomarker data, such as B-type natriuretic peptide, were unavailable in this study. Consequently, stage C HF was classified solely on the basis of current or previous symptoms of HF together with evidence of structural or functional cardiac abnormalities identified by echocardiography. The generalizability may also be limited. While this is a representative population-based study recruited without regard to health status, there are certainly portions of the population that may not have volunteered for the study, including those with other serious medical conditions. As such, this may reflect healthier than average population in Tobago. Further studies will be need to determine the extent to which these results generalize to other African Caribbean nations or to persons from African Caribbean nations living in different regions such as immigrants to the U.S. or U.K, which both have high representation from the African Caribbean diaspora ^43^.

## Conclusions

Cardiac remodeling in community-dwelling African Caribbeans aged 40+ years in Tobago is common, present in the majority of individuals. Female and older individuals have more DD compared to males and younger people. Additionally, the older population tended to have slightly more concentric remodeling and LVH than the younger population. Women also have a higher prevalence of early-stage HF (Stages A and B), whereas men are more likely to have symptomatic (Stage C) HF. These findings may not only inform public health priorities but also serve as a reference for future research, including longitudinal studies, to investigate predictors of the abnormal cardiac structure and function among these African Caribbeans in Tobago. In future studies, we aim to evaluate the progression of concentric remodeling and its potential influence through longitudinal follow-up in these African Caribbean adults.

## Data Availability

All data produced in the present study are available upon reasonable request to the authors

---

**Supplementary Table 1:**
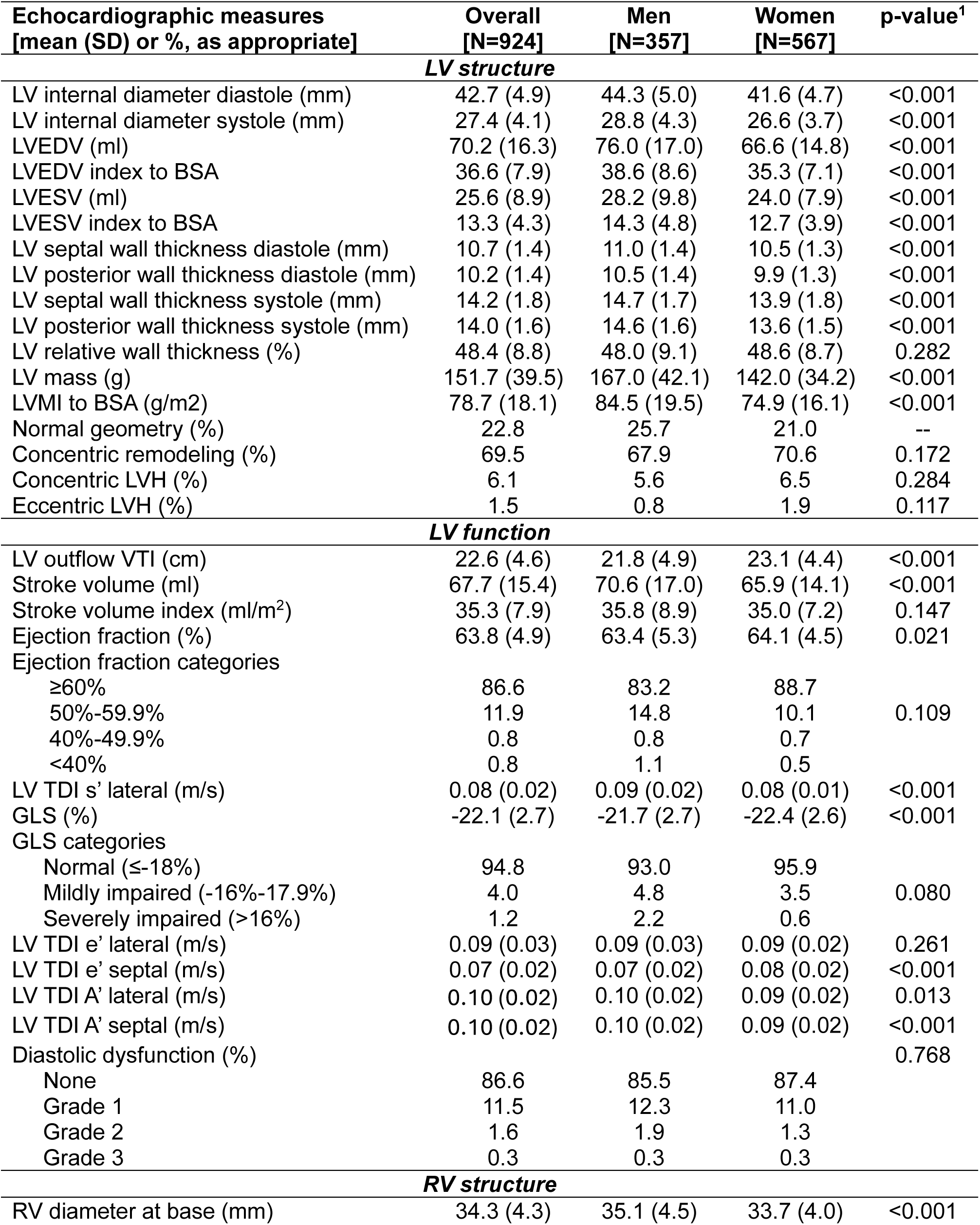
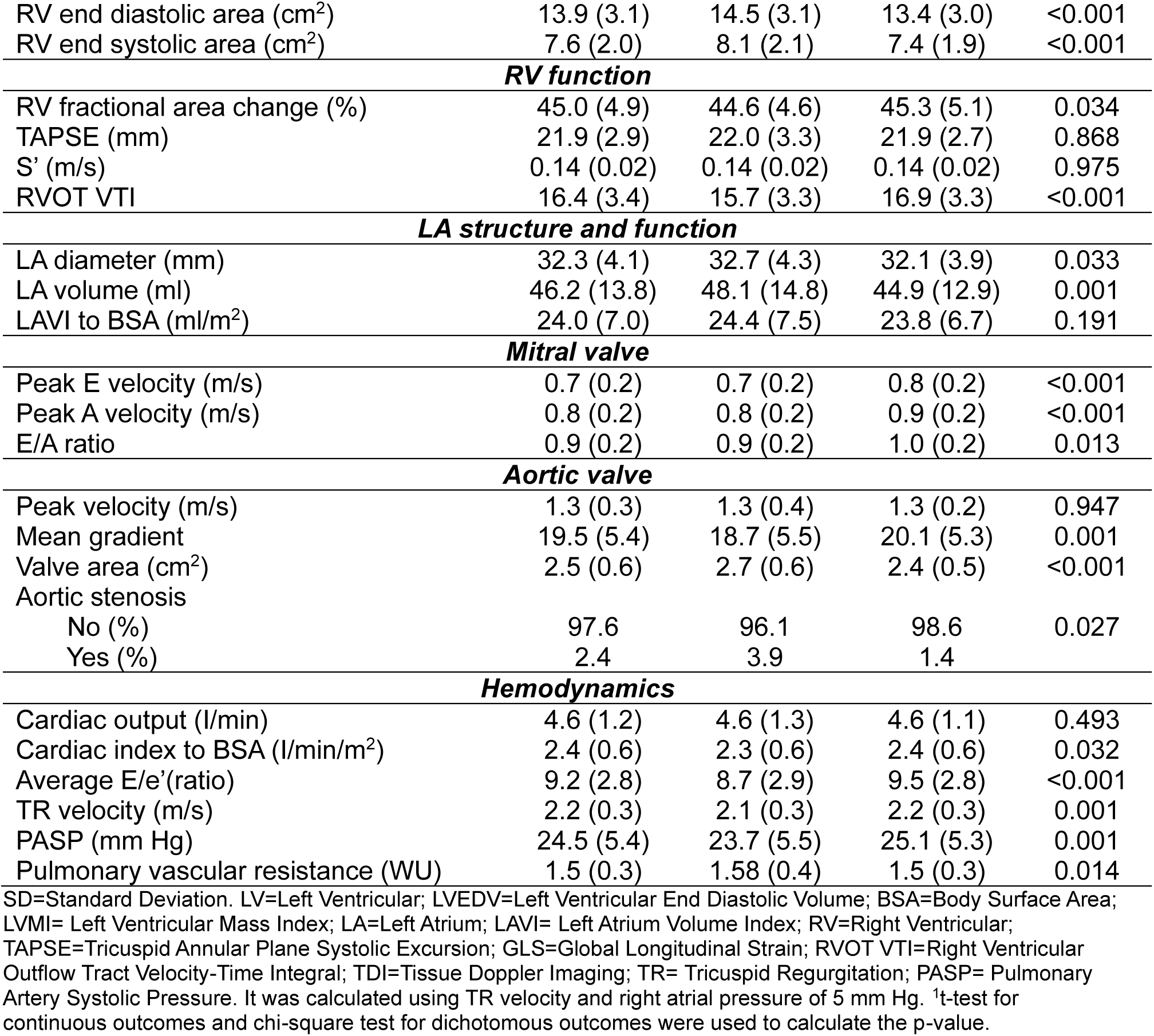
Sex specific comparisons of cardiac structure and function in the THS.

